# Prevalence, Genetics, and Imaging Characteristics of Patients with Mitral Valve Prolapse and Arrhythmogenic Right Ventricular Cardiomyopathy

**DOI:** 10.64898/2026.05.14.26353246

**Authors:** Amy Rich, Lionel Tastet, Luca Cristin, Rohit Jhawar, Janet J. Tang, Melvin Scheinman, Francesca N. Delling

## Abstract

**Background:** Concomitant arrhythmogenic right ventricular cardiomyopathy (ARVC) and mitral valve prolapse (MVP) has only been described in case reports. Little is known about genetic and phenotypic characteristics of these patients.

**Objective:** To describe the prevalence, genetics, and imaging characteristics of MVP in ARVC patients.

**Methods:** We identified 111 definite ARVC cases through medical record review, arrhythmia/cardiomyopathy targeted gene panels, and contrast cardiac magnetic resonance data. MVP was diagnosed on echocardiography as mitral leaflet displacement >2 mm above the annular plane in systole, “borderline MVP” defined as ≤2 mm.

**Results:** We found MVP/borderline MVP in 14% of ARVC patients. Cardiac arrest occurred in 20% of those with MVP/borderline MVP compared to 16% without valve abnormalities. Among 69 ARVC patients with identified genetic variants, *PKP2* mutations were highly prevalent (64%), particularly in those with MVP (83%). Most MVPs had posterior prolapse (73%) and trace/mild mitral regurgitation (87%). None had mitral annular disjunction. ARVCs with MVP had higher LV mass (93 vs. 75 g/m^2^, p = 0.02) and a higher prevalence of LV wall motion abnormalities (27% vs. 5%, p = 0.02) compared to ARVCs without valve abnormalities.

**Conclusions:** MVP is prevalent in ARVC and characterized by *PKP2* variants in most cases. Typical features of arrhythmic MVP like bileaflet involvement and annular disjunction are rare in ARVC with MVP; features of arrhythmogenic left-sided cardiomyopathy – increased LV mass index and wall motion abnormalities – are more common. Further studies are needed to understand the role of MVP in arrhythmic risk stratification of ARVC.

**Key Points:**

1. In a cohort of 111 ARVC patients, 14% had MVP.
2. Among 69 concomitant MVP and ARVC patients with identified genetic variants, *PKP2* mutations were highly prevalent (83%).
3. Typical features of arrhythmic MVP like bileaflet involvement and annular disjunction are rare in ARVC with MVP, while features of arrhythmogenic left-sided are more common.
4. Further studies are needed to understand the role of *PKP2* genetic variation and MVP interaction in arrhythmic risk stratification of and left ventricular involvement in ARVC.

## INTRODUCTION

Mitral valve prolapse (MVP) is a common valve disorder affecting approximately 2-3% of individuals in the general population.^1, 2^ MVP is the leading cause of significant mitral regurgitation (MR) requiring surgical intervention.^3^ Although MVP has traditionally been considered benign in the absence of severe MR,^4–6^ the yearly incidence of sudden cardiac death or arrest can range between 0.14 and 1.8%.^2, 7–9^ Established risk factors for ventricular arrhythmias in MVP are young age, female sex, bileaflet involvement, mitral annular disjunction (MAD), frequent or complex ventricular ectopy, and myocardial fibrosis attributable to excess leaflet motion.^2, 10–13^ Ventricular arrhythmias in MVP are thought to be the consequence of abnormal, localized valvular-myocardial mechanics.^14^ However, a diffuse primary myopathic process has also been proposed by our group and others^15,16^ and corroborated by the discovery of cardiomyopathy genes in large genome wide association studies of MVP.^17^

Arrhythmogenic Right Ventricular Cardiomyopathy (ARVC) is primarily a genetic condition and is characterized by fibrofatty infiltration of the right ventricle.^18^ ARVC is found in 1 in every 2,000-5,000 individuals in the general population.^18^ In early stages, structural changes may be absent or subtle, and individuals are often asymptomatic but at risk for sudden cardiac death, particularly during exertion.^19^ Progression to more advanced right ventricular (RV) disease occurs in later stages, and can manifest as arrhythmogenic syncope, sustained ventricular tachycardia (VT) secondary to the formation of scar, and cardiac arrest or sudden cardiac death.^20^ Later stages also involve more clearly discernible RV morphological abnormalities - notably dilated cardiomyopathy – identified through echocardiographic and cardiac magnetic resonance (CMR) imaging.^20^ Genetic mutations in desmosomal genes, including

*PKP2*, *DSP*, *DSG2*, *DSC2*, and *JUP,* and non-desmosomal genes including *TMEM43, DES,* and *PLN,* act as fundamental players in muscle contraction synchronization and account for the majority of pathogenic or likely pathogenic variants in ARVC.^18, 21^

ARVC has been historically characterized as a dominant RV cardiomyopathy; however, more recent research has described the majority of ARVC cases to involve biventricular structural and functional changes, independent of age at death, sex, macroscopically normal appearance of the heart at autopsy, or competitive sport participation.^19, 22^ Left-dominant arrhythmogenic cardiomyopathy is characterized by a proneness to ventricular arrhythmias in early disease stages that is disproportionate to observed structural and systolic function abnormalities.^23^ It is proposed that LV involvement in ARVC may be more prevalent in patients with *PKP2, DSP, DSG2,* or *TMEM43* gene variants, which have been associated with LV late gadolinium enhancement and chamber dilatation.^18,22,24^

Our understanding of left-sided cardiomyopathy in ARVC is limited, especially regarding left-sided concomitant diseases in ARVC. The co-existence of MVP and ARVC has only been described in case reports^25–27^; however, little is known about the genetic and phenotypic features of patients with concomitant ARVC and MVP, especially in the setting of a left-sided cardiomyopathy. In this study, we describe the genetics and imaging characteristics in a cohort of ARVC patients with MVP. We hypothesized that features of arrhythmogenic left-sided cardiomyopathy will be more prevalent in the MVP cohort.

## METHODS

### Study Population

We searched the University of California, San Francisco (UCSF) electronic medical records and the UCSF Cardiovascular Genetics Center Database for patients with a diagnosis of ARVC based on an International Classification of Disease code between 2015 and 2025. Patients were included in our study if they presented a definite ARVC diagnosis based on the 2010 revised ARVC diagnostic Task Force Criteria.^19^ Definite ARVC Diagnostic terminology for revised 2010 Task Force Criteria are described in detail in the **Supplemental Material**. We corroborated these diagnoses by reviewing arrhythmia/cardiomyopathy targeted whole exome sequencing panels, clinical notes and CMR data. The study population included individuals involved in regular sports activity prior to ARVC diagnosis. In these subjects, CMR reference cut-offs for ventricular dimensions were similar to the ones used for the remaining study sample.

### Cardiovascular Genetics

Genetic variant information was collected through review of arrhythmia/cardiomyopathy targeted clinical whole exome sequencing panels in the UCSF electronic medical records and the UCSF Cardiovascular Genetics Center Database. Genetic variant details are reported in the **Supplemental Material**.

### Clinical Data

We collected the following clinical data from the electronic health records: age at the time of each patient’s most recent echocardiogram, race, ethnicity, history of smoking, hypertension or diabetes, history of cardiac arrest treated with an implantable cardioverter-defibrillator (ICD), history of ventricular ectopy, and family history of cardiac arrest.

### Echocardiographic and CMR assessment

Two-dimensional echocardiograms were performed using commercially available ultrasound systems. Details of the CMR acquisition and image analysis are reported in the **Supplemental Material**.

#### ARVC

CMR was utilized to assess RV and LV wall motion abnormalities (dyskinesis, akinesis, hypokinesis), RV ejection fraction, presence of late gadolinium enhancement in the LV and/or RV myocardium, and the presence of diffuse fatty infiltration. CMR was also utilized to measure RV end-diastolic/end-systolic volumes, which were indexed to body surface area. CMR criteria for ARVC included the presence of RV dyskinesis/akinesis and either RV dilatation or systolic dysfunction.^19^ LV involvement in ARVC (“left ventricular arrhythmogenic cardiomyopathy”) was defined on CMR as presence of any left ventricular structural or functional abnormalities, including LV enlargement; LV dyskinesis, akinesis, or hypokinesis; late gadolinium enhancement in the LV; and fatty infiltration in the LV.

#### MVP

MVP was diagnosed on both echocardiography and CMR as mitral leaflet displacement > 2 mm above the annular plane in systole^28^ in a parasternal long-axis or 3-chamber view. MVP subtypes were categorized as bileaflet or single leaflet prolapse (anterior prolapse, posterior prolapse). Participants with ≤2 mm nondiagnostic mitral leaflet displacement were included in the analysis as “borderline MVP.”^29^ We have previously demonstrated^30,31^ that these borderline morphologies resemble MVP genetically and phenotypically and can progress to fully diagnostic MVP.^32^ All MVP diagnoses from echocardiographic reports were corroborated by independent image review (F.N.D. and L.C.). Echocardiographic images were also reviewed for the presence of MAD, a separation between the left atrial/mitral leaflet junction and the basal inferolateral LV myocardium.^33^ CMR long-axis images were analyzed to identify lesser degrees of MAD that may have been missed on echocardiography. On both imaging modalities, care was taken to distinguish true MAD from pseudo-MAD, where the posterior mitral leaflet curls on itself creating a false impression of MAD.^34^ Curling was visually assessed in the parasternal long-axis view at end-systole by an independent echocardiographer (L.C.) and was defined as abnormal inward and apical motion of the basal infero-lateral wall associated with annular expansion.^2^

### Other echocardiographic and CMR variables

MR, aortic regurgitation, and tricuspid regurgitation severity were assessed on echocardiographic images using a multiparametric approach, as recommended by guidelines,^35^ and graded as: none, trace, mild, moderate, or severe. LV end-diastolic/end-systolic volumes and LV mass were indexed to body surface area. Other echocardiographic measurements included LV ejection fraction, RV dimensions and systolic function (semi-quantitative assessment) and pulmonary artery systolic pressure. CMR was used to calculate indexed RV volumes and RV ejection fraction.

### Statistical Methods

The study design is retrospective and patients were included consecutively. Missing data were excluded. Continuous variables were expressed as a mean ± SD or median (interquartile range [IQR]) and were tested for equality of variances with the Folded F-test. Continuous variables were compared between groups using Welch’s t-test or a Student’s t-test as appropriate. Categorical variables were presented as frequencies and percentages and were compared with Fisher’s Exact Test. All p-values were two-sided, and statistical significance was set at less than 0.05. Statistical analyses were performed using SAS 9.4 (SAS Institute, Cary, NC).

## RESULTS

### Study population characteristics

The study population comprised 111 definite ARVC cases, of which 60 were women and 51 were men. The average age at the time of each patient’s most recent echocardiogram was 47 years old, with race breakdown shown in **Table 1**. The majority (77%) of patients had a primary or secondary prevention ICD, 17% had a cardiac arrest, 82% had premature ventricular contractions, 28% had non-sustained VT, 62% had sustained VT, and 19% had hypertension. Among the 111 definite ARVC patients, we identified 15 MVP cases. Clinical and echocardiographic characteristics of the study population are presented in **Table 1**. **Figure 1A** illustrates an example of a definite ARVC case with posterior MVP on an echocardiographic parasternal long-axis view. **Figure 1B** shows biventricular involvement and areas of free wall RV dyskinesis in the same ARVC patient on a CMR 4-chamber view.

**Table 1.** Values are mean (SD), median (IQR), or absolute number (%). ICD = implantable cardioverter defibrillator; LVEDVI = left ventricular end-diastolic volume index; LVEDSI = left ventricular end-systolic volume index; MAD = mitral annular disjunction; MVP = mitral valve prolapse; NSVT = non-sustained ventricular tachycardia; *PKP2* = plakophilin-2; PVC = premature ventricular contractions; RVEDVI = right ventricular end-diastolic volume index; RVESVI = right ventricular end-systolic volume index; VT = sustained ventricular tachycardia.

| **Table 1** | **All ARVC**<br>**patients**<br>**n = 111** | **MVP**<br>**n = 15** | **Non-MVP**<br>**n = 96** | **p-value** |
| --- | --- | --- | --- | --- |
| **Clinical characteristics** |  |  |  |  |
| Age, y | 47 ± 15 | 52 ± 14 | 46 ± 15 | 0.16 |
| Sex, n (%) |  |  |  | 0.27 |
| Male | 51 (46) | 9 (60) | 42 (44) |  |
| Female | 60 (54) | 6 (40) | 54 (56) |  |
| Race, n (%) |  |  |  | 0.47 |
| White | 70 (64) | 11 (73) | 59 (62) |  |
| Black | 4 (4) | 1 (7) | 3 (3) |  |
| Black+White | 1 (1) | 0 (0) | 1 (1) |  |
| Asian | 17 (15) | 1 (7) | 16 (17) |  |
| Asian+White | 3 (3) | 1 (7) | 2 (2) |  |
| Unknown | 16 (14) | 1 (7) | 15 (16) |  |
| ICD, n (%) | 85 (77) | 11 (73) | 74 (77) | 0.75 |
| Cardiac Arrest, n (%) | 19 (17) | 3 (20) | 16 (17) | 0.72 |
| Family History of Cardiac Arrest, n (%) | 43 (39) | 6 (40) | 37 (39) | 1.00 |
| PVCs, n (%), | 91 (82) | 11 (73) | 80 (83) | 0.47 |
| NSVT, n (%) | 31 (28) | 5 (33) | 26 (27) | 0.76 |
| VT, n (%) | 69 (62) | 3 (87) | 66 (69) | 0.22 |
| Smoking, n (%) | 17 (15) | 3 (20) | 14 (15) | 0.70 |
| Hypertension, n (%) | 21 (19) | 4 (27) | 17 (18) | 0.48 |
| Diabetes, n (%) | 6 (5) | 0 (0) | 6 (6) | 1.00 |
| **Clinical Genetics Characteristics** |  |  |  |  |
| Patients with available genetic testing, n (%) | 90 (81) | 10 (11*) | 80 (89*) | 0.47 |
| Patients with at least one identified genetic variant, n (%) | 69 (62) | 6 (60*) | 63 (79*) | 0.13 |
| Patients with genetic variation in *PKP2*, n (%) | 44 (40) | 5 (83*) | 39 (62*) | 1.00 |
| **Echocardiographic characteristics** |  |  |  |  |
| MVP anatomy, n (%) |  |  |  |  |
| Anterior | 3 (3) | 3 (20) |  |  |
| Posterior | 11 (10) | 11 (73) |  |  |
| Bileaflet | 1 (1) | 1 (7) |  |  |
| Curling, n (%) | 4 (4) | 4 (27*) |  |  |
| MAD, n (%) | 0 (0) | 0 (0) |  |  |
| Tricuspid Valve Prolapse, n (%) | 0 (0) | 0 (0) |  |  |
| Left ventricular ejection fraction, % | 57 ± 11 | 56 ± 7 | 58 ± 11 | 0.43 |
| LVEDVI, mL/m<sup>2</sup> | 49.55 ± 21.98 | 52.22 ± 13.92 | 49.07 ± 23.16 | 0.64 |
| LVESVI, mL/m<sup>2</sup> | 20.83 ± 16.11 | 23.37 ± 9.76 | 20.37 ± 17.01 | 0.38 |
| Left Ventricular Mass, g/m<sup>2</sup> | 77 ± 24 | 93 ± 24 | 74 ± 23 | **0.02** |
| Right Ventricular Size, n (%) |  |  |  | 0.36 |
| Normal | 29 (26) | 1 (17*) | 28 (36*) |  |
| Mildly increased | 16 (14) | 1 (17*) | 15 (19*) |  |
| Moderately increased | 22 (20) | 1 (17*) | 21 (27*) |  |
| Severely increased | 17 (15) | 3 (50*) | 14 (18*) |  |
| Right Ventricular Function, n (%) |  |  |  | 0.73 |
| Normal | 39 (35) | 4 (27) | 35 (41*) |  |
| Mild dysfunction | 28 (25) | 5 (33) | 23 (27*) |  |
| Moderate dysfunction | 21 (19) | 4 (27) | 17 (20*) |  |
| Severe dysfunction | 13 (12) | 2 (13) | 11 (13*) |  |
| Pulmonary Artery Systolic Pressure, mmHg | 26 ± 6 | 26 ± 6 | 26 ± 6 | 0.89 |
| Mitral regurgitation grade, n (%) |  |  |  | **0.01** |
| None | 24 (22) | 0 (0) | 24 (31*) |  |
| Trace | 50 (45) | 9 (75*) | 41 (53*) |  |
| Mild | 12 (11) | 1 (8*) | 11 (14*) |  |
| Moderate | 3 (3) | 2 (17*) | 1 (1*) |  |
| Severe | 0 (0) | 0 (0) | 0 (0) |  |
| Tricuspid regurgitation grade, n (%) |  |  |  | 0.42 |
| None | 6 (5) | 0 (0) | 6 (7*) |  |
| Trace | 28 (25) | 3 (21*) | 25 (31*) |  |
| Mild | 29 (26) | 7 (50*) | 22 (27*) |  |
| Moderate | 23 (21) | 2 (14*) | 21 (26*) |  |
| Severe | 10 (9) | 2 (14*) | 8 (10*) |  |
| Aortic regurgitation grade, n (%) |  |  |  | 0.33 |
| None | 63 (57) | 6 (68*) | 57 (83*) |  |
| Trace | 5 (5) | 1 (11*) | 4 (6*) |  |
| Mild | 9 (8) | 2 (22*) | 7 (10*) |  |
| Moderate | 0 (0) | 0 (0) | 0 (0) |  |
| Severe | 1 (1) | 0 (0) | 1 (1) |  |
| **Cardiovascular Magnetic Resonance Characteristics** |  |  |  |  |
| Right ventricular wall motion abnormalities, n (%) | 49 (44) | 9 (60) | 40 (42) | 0.26 |
| RV Outflow tract | 7 (14) | 2 (22) | 5 (13) |  |
| Free wall (basal, mid, apical) | 33 (67) | 7 (78) | 26 (65) |  |
| RV Apex | 5 (10) | 1 (11) | 4 (10) |  |
| Global motion abnormalities | 15 (31) | 3 (33) | 12 (30) |  |
| Left Ventricular wall motion abnormalities, n (%) | 9 (8) | 4 (27) | 5 (5) | **0.02** |
| Basal inferior | 1 (11) | 1 (25) | 0 (0) |  |
| Mid anterior wall | 2 (22) | 1 (25) | 1 (20) |  |
| Mid anteroseptal wall | 1 (11) | 0 (0) | 1 (20) |  |
| Mid inferoseptal wall | 1 (11) | 0 (0) | 1 (20) |  |
| Mid inferolateral wall | 2 (22) | 1 (25) | 1 (20) |  |
| Apical anterior | 1 (11) | 0 (0) | 1 (20) |  |
| Apical septal | 1 (11) | 0 (0) | 1 (20) |  |
| Apical inferior | 2 (22) | 1 (25) | 1 (20) |  |
| Apical lateral | 2 (22) | 1 (25) | 1 (20) |  |
| Global motion abnormalities | 3 (33) | 1 (25) | 3 (60) |  |
| Right Ventricular Ejection Fraction, % | 36 ± 19 | 36 ± 6 | 42 ± 12 | 0.24 |
| RVEDVI, mL/m<sup>2</sup> | 109.77 ± 35.95 | 117.87 ± 24.98 | 108.77 ± 37.76 | 0.55 |
| RVESVI, mL/m<sup>2</sup> | 66.46 ± 32.34 | 70.6 ± 16.84 | 65.95 ± 33.85 | 0.77 |
| Diffuse fatty infiltration, n (%) | 19 (17) | 6 (40) | 13 (14) | **0.02** |
| RV free wall | 13 (68) | 4 (67) | 9 (69) |  |
| RVOT | 3 (16) | 0 (0) | 3 (23) |  |
| Left apical lateral wall | 1 (5) | 0 (0) | 1 (8) |  |
| Left apical septal wall | 1 (5) | 0 (0) | 1 (8) |  |
| Septal wall | 2 (11) | 1 (17) | 1 (8) |  |
| Late Gadolinium Enhancement in Right Ventricle, n (%) | 21 (19) | 2 (29*) | 19 (42*) | 0.69 |
| Late Gadolinium Enhancement in Left Ventricle, n (%) | 16 (14) | 3 (33*) | 13 (34*) | 1.00 |

\*prevalence based on available clinical, genetic and imaging data

### ARVC patients with MVP

#### Clinical characteristics

The prevalence of fully diagnostic or borderline MVP among ARVC patients was 14% (n=15, including 7 MVP) (mean age 52±15 years; 40% females). Seventy-three% (n=11) of MVPs within the ARVC cohort had an ICD, 20% (n=3) had a history of smoking, 27% (n=4) had hypertension, and none had diabetes. Forty% (n=6) had a family history of cardiac arrest.

#### Genetic characteristics

Among 69 ARVC patients with identified genetic variants, *PKP2* genetic variation was highly prevalent (64%). Those with MVP/borderline MVP in particular demonstrated either a *PKP2* pathogenic variant (n=4) or a variant of uncertain significance (VUS) (n=1) (excluding likely benign VUS) in 5/6 or 83% of cases. *PKP2* and non-*PKP2* genetic variants are listed in the **Supplemental table**.

#### Echocardiographic Characteristics

Posterior prolapse was the most common mitral valve anatomy among MVPs (11/15 or 73%), while 20% (n=3) had anterior MVP and a small minority (7% or n=1) had bileaflet prolapse. Of the 13 MVPs in the ARVC cohort with interpretable color Doppler, most had no/trace MR (77% or n=10), while 1 had mild MR (8.3%), 2 had moderate MR (16.7%), and 0 had severe MR. None had mitral annular disjunction. Four MVPs had curling (27%). The average LV ejection fraction in the MVP cohort was 56%. The average LVEDVi and LVESVi were 52 ml/m^2^ and 23 ml/m^2^, respectively. The average LV mass, indexed to body surface areas, was 93 g/m^2^. Among the 6 MVPs with available RV ventricular volume data, 17% (n=1) had normal volumes, 33% (n=2) had mildly or moderately increased volume, and 50% (n=3) had severely increased volume. Four of the 15 MVPs (27%) had normal RV function, while 33% (n=5) had mild, 27% (n=4) had moderate, and 13% (n=2) had severe RV dysfunction, respectively. The average pulmonary artery systolic pressure in the MVP group was 26 mmHg. Most ARVC patients with MVP had mild or moderate tricuspid regurgitation (TR) **(Table 1)**. None had significant aortic regurgitation.

#### CMR Characteristics

Seven of the ARVC patients with MVP/borderline MVP who underwent CMR had available RV late gadolinium enhancement data, and 29% (n=2/7) had evidence of late gadolinium enhancement in the RV myocardium. Nine of the MVPs had available LV late gadolinium enhancement data. One third (n=3/9) had evidence of late gadolinium enhancement in the LV myocardium. Sixty% (n=9/15) of the MVP group had RV wall motion abnormalities, and 27% (n=4/15) had LV wall motion abnormalities. The average RV ejection fraction, right ventricular end-diastolic volume index, and right ventricular end-systolic volume index are reported in **Table 1**. Six cases out of the 15 MVPs (40%) had evidence of diffuse fatty infiltration of the RV. None had evidence of diffuse fatty infiltration of the LV. The most notable echocardiographic and CMR findings in the MVP subgroup are summarized in **Figure 2**.

### Comparative Analysis between ARVC with and without MVP

Our comparative analysis between the MVP and non-MVP groups demonstrated greater biventricular involvement in concomitant ARVC and MVP. Specifically, ARVC patients with MVP had statistically significant higher average LV mass indexed to body surface areas compared to ARVC patients with no valve abnormalities (93 g/m^2^vs. 75 g/m^2^, p = 0.02) (**Figure 3).** There was a higher prevalence of LV wall motion abnormalities in the MVP group compared to the non-MVP group (27% vs. 5%). This difference was significant at a p-value of 0.02. Genetics and imaging characteristics and comparative analyses excluding borderline MVP cases are reported in the **Supplemental Material.**

### Arrhythmic Outcomes between ARVC with and without MVP

Three of 15 or 20% of ARVC cases with MVP/borderline MVP had a cardiac arrest compared to 15/96 or 16% of individuals with ARVC but no valve abnormalities. There was a lower prevalence of premature ventricular contractions (73% (n= 11) among ARVC patients with MVP compared to those without MVP (83% (n=80). However, a higher percentage of ARVC cases with MVPs (33%) (n=5) had non-sustained VT compared to 27% (n=26) of the non-MVP ARVC group **(Table 1)**. Similarly, 87% (n=13) of the MVP group had sustained VT, compared to only 69% (n=66) in the ARVC group with no valve abnormalities **(Table 1)**.

## DISCUSSION

In this cohort study of ARVC patients, we describe the prevalence of MVP, and the genetic, clinical and imaging characteristics of patients with concomitant MVP. *PKP2* genetic variation was a prevalent finding of MVP with ARVC. We observed that typical features of arrhythmic MVP such as bileaflet involvement and annular disjunction are rare in MVPs with ARVC, whereas features of arrhythmogenic left-sided cardiomyopathy – increased LV mass indexed to body surface area and LV wall motion abnormalities – are more common.

### Prevalence of MVP among ARVC patients

The overall prevalence of combined MVP and borderline MVP in our ARVC sample was 14% (7% for borderline MVP alone), higher than 2.5% (0.3% for borderline MVP alone) in the general population. ^4, 31^ These findings suggest that the co-existence of mitral valve abnormalities and ARVC may be genetically determined rather than coincidental. It is plausible that *PKP2* mutations that result in desmosome dysfunction may serve as a substrate for myocardial fibrofatty degeneration and mitral leaflet structural abnormalities, manifesting clinically as concurrent ARVC and MVP. Further studies are needed to corroborate this hypothesis.

### PKP2 in concomitant MVP and ARVC

Among patients in our study with concomitant MVP and ARVC, 73% had posterior prolapse and 83% had pathogenic variants or variants of unknown significance (VUS) in the *PKP2* gene. Of importance, previous case reports have described sudden cardiac arrest, sudden cardiac death, and heart failure occurring in *PKP2* mutation carriers more frequently than in those with other genetic mutations.^36,37^ We also observed a statistically significant higher prevalence of overall diffuse fatty infiltration of the RV and the LV in the MVP group compared to the non-MVP group (40% vs. 14%, p = 0.02), which may be explained by the prominent role of *PKP2* in desmosmal dysfunction and in fatty infiltration. The high prevalence of *PKP2* genetic variation among those with MVP and ARVC warrants future screening of ARVC patients for MVP, especially posterior mitral valve prolapse, to better stratify risk of ventricular arrhythmias and sudden cardiac events in ARVC patients.

### Increased left ventricular mass index in concomitant MVP and ARVC

In our study, MVP patients had higher LV mass indexed to body surface area compared to the non-MVP group. Of note, 87% of the MVP patients had no or trace/mild MR, which suggests a different underlying etiology for increased LV mass in ARVC with MVP not attributable to MR.^38^ While increased LV mass in MVP may be a consequence of localized hypertrophy in the basal inferolateral wall due to traction by the prolapsing leaflets in bileaflet MVP/ MAD,^33^ a primary, genetic cardiomyopathy may be more relevant in our sample of MVPs with mostly posterior MVP and no MAD. A diffuse myopathic process had been previously described by our group and others in MVP patients.^15,39,40^ Moreover, within the spectrum of arrhythmogenic cardiomyopathies, desmosome dysfunction can lead to significant LV structural changes, including fibrotic remodeling and increased LV mass in both mouse models and human studies.^41, 42^ It is well-established that LV hypertrophy is associated with increased risk for ventricular arrhythmias,^43–44^ regardless of the cause.

### Prevalence of left-ventricular wall motion abnormalities in concomitant MVP and ARVC

We found a statistically significant higher prevalence of LV wall motion abnormalities in the group of ARVC patients with MVP compared to those with normal mitral valves. The locations of segmental wall motion abnormalities are detailed in **Table 1**. None of the patients had coronary artery disease. Inferolateral wall hypokinesis may be the consequence of repetitive traction by the prolapsing leaflets and MAD with development of replacement fibrosis in the same area.^14^ However, in our study, no MVPs or borderline MVPs had MAD, only 27% had curling, and only 33% of the MVPs with available contrast CMR data had LV myocardial late gadolinium enhancement. Hence, LV wall motion abnormalities, especially when not involving the inferolateral wall, cannot be explained by localized traction and may be the consequence of a diffuse myopathic/multifocal process involving both the LV and RV.^45^ The presence of segmental LV wall motion abnormalities in ARVC is correlated with a higher risk of ventricular arrhythmias,^46^ although prior studies have failed to investigate the influence of MVP on this ARVC subtype.

### Study Strengths and Weaknesses

The retrospective nature of this study precludes causal inference. Additionally, the small sample size of concomitant MVP and ARVC reduces the power of more advanced statistical analyses of MVP and LV involvement in ARVC. Ideally, only pathogenic variants should have been included in our analysis. However, because of the small size of our ARVC cohort with available genetic data, we opted to include VUS in the *PKP2* gene to the pool of analyzed variants based on the fact that none were notated in additional analyses as “VUS likely benign” and that they represent variants within a gene that has been linked to ARVC in multiple studies.^21,47,48^ An additional limitation relates to our inclusion of “borderline MVP” within the primary MVP cohort. Prior work has shown that borderline morphologies may share genetic and phenotypic features with fully diagnostic MVP and can progress over time.^30, 31^ Despite these findings, only approximately one-third of individuals with borderline MVP in the Framingham Heart Study progressed to fully diagnostic MVP over longitudinal follow-up, and cardiovascular outcomes among individuals with borderline MVP were comparable to those of referent participants, whereas patients with true MVP demonstrated a trend toward greater cardiovascular events.^32^ Therefore, in some patients, borderline MVP may represent a heterogeneous or lower-risk phenotype rather than simply an early stage of classic MVP. Grouping borderline and fully diagnostic MVP together may have introduced phenotypic heterogeneity and could have influenced our estimates of prevalence. However, we provide separate analyses excluding borderline MVP cases in the Supplemental Material to partially address this limitation. Finally, the ARVC cohort was assembled from a tertiary referral center, which likely introduced selection bias toward more advanced and arrhythmically active disease, as evidenced by the high rates of ICD implantation and RV dysfunction. The observed prevalence of MVP in this cohort should therefore be interpreted with caution, as it may not accurately represent the prevalence of MVP across the full spectrum of ARVC, including early-stage or genotype-positive/phenotype-negative patients.

Despite these limitations, our study offers a sufficiently large sample size considering the rarity of ARVC. Importantly, our study is the first to provide a comprehensive overview of genetic, clinical, and imaging characteristics of concomitant MVP and ARVC. Our study is also the first to find a statistically significant higher average indexed LV mass and statistically significant higher prevalence of LV wall motion abnormalities in a sample with concomitant MVP and ARVC compared to ARVC patients with no valve abnormalities. Prospective, longitudinal studies are needed to investigate the interaction of *PKP2* genetic variation and MVP in risk stratification of ARVC. In addition, mechanistic studies are needed to better understand the underlying etiology and myocardial mechanisms of MVP that may serve as a substrate for left ventricular involvement and left ventricular arrhythmias in ARVC.

## CONCLUSIONS

MVP is a prevalent finding in ARVC and is characterized by *PKP2* genetic variation in most cases. Typical features of arrhythmic MVP such as bileaflet involvement and annular disjunction are rare in MVPs with ARVC, whereas features of arrhythmogenic left-sided cardiomyopathy are more common. Further studies are needed to understand the role of concomitant MVP in arrhythmic risk stratification of ARVC patients.

## Data Availability

All data produced in the present study are available to University of California, San Francisco affiliated researchers.

## Statements and Declarations

Not applicable

## Ethical Considerations

The study was conducted in accordance with the Declaration of Helsinki and was approved by the University of California, San Francisco Institutional Review Board (no. 17-22679) on October 07, 2010, with the need for written informed consent waived.

## ABBREVIATIONS

ARVC: Arrhythmogenic Right Ventricular Cardiomyopathy
CMR: Cardiovascular Magnetic Resonance
ICD: Implantable Cardioverter Defibrillator
LV: left ventricular
MVP: mitral valve prolapse
MR: mitral regurgitation
MAD: mitral annular disjunction
*PKP2*: Plakophilin 2
RV: right ventricular
VT: ventricular tachycardia

---

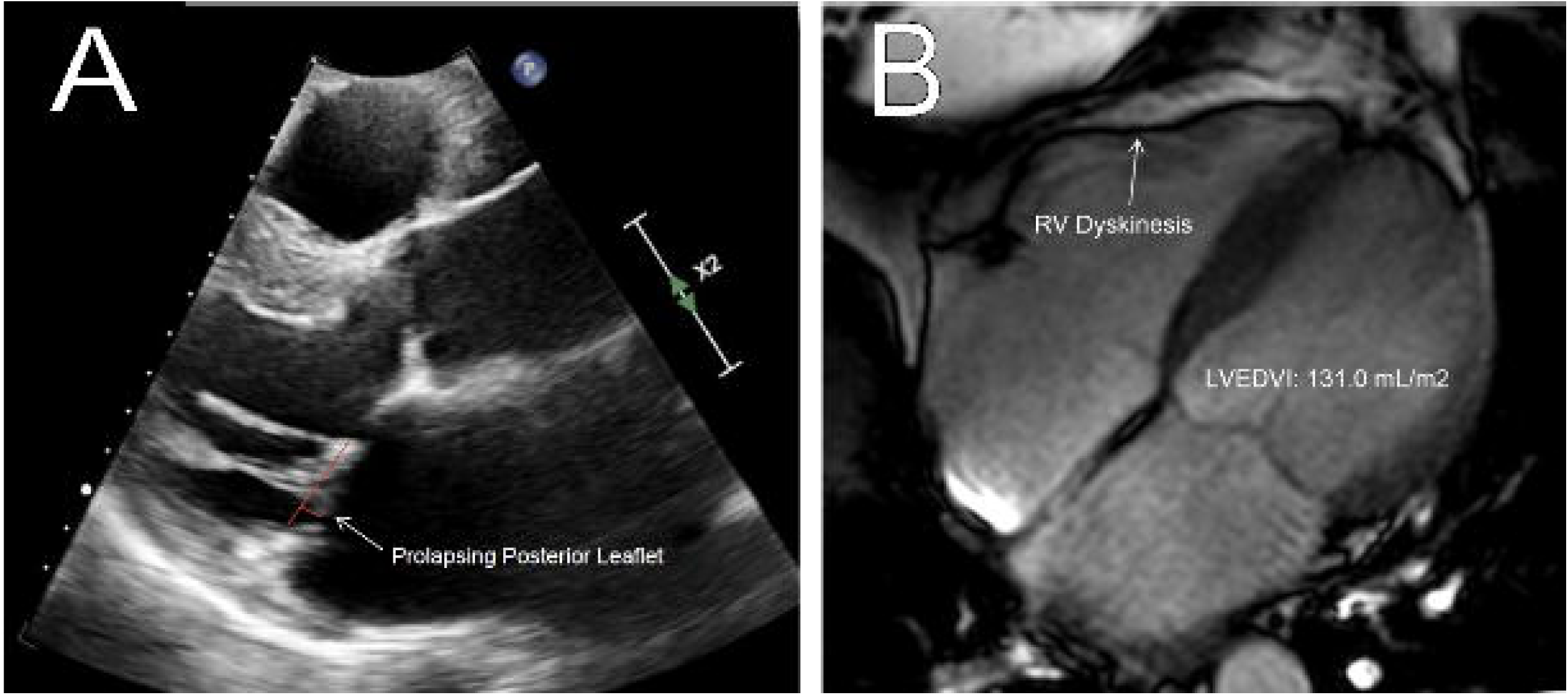

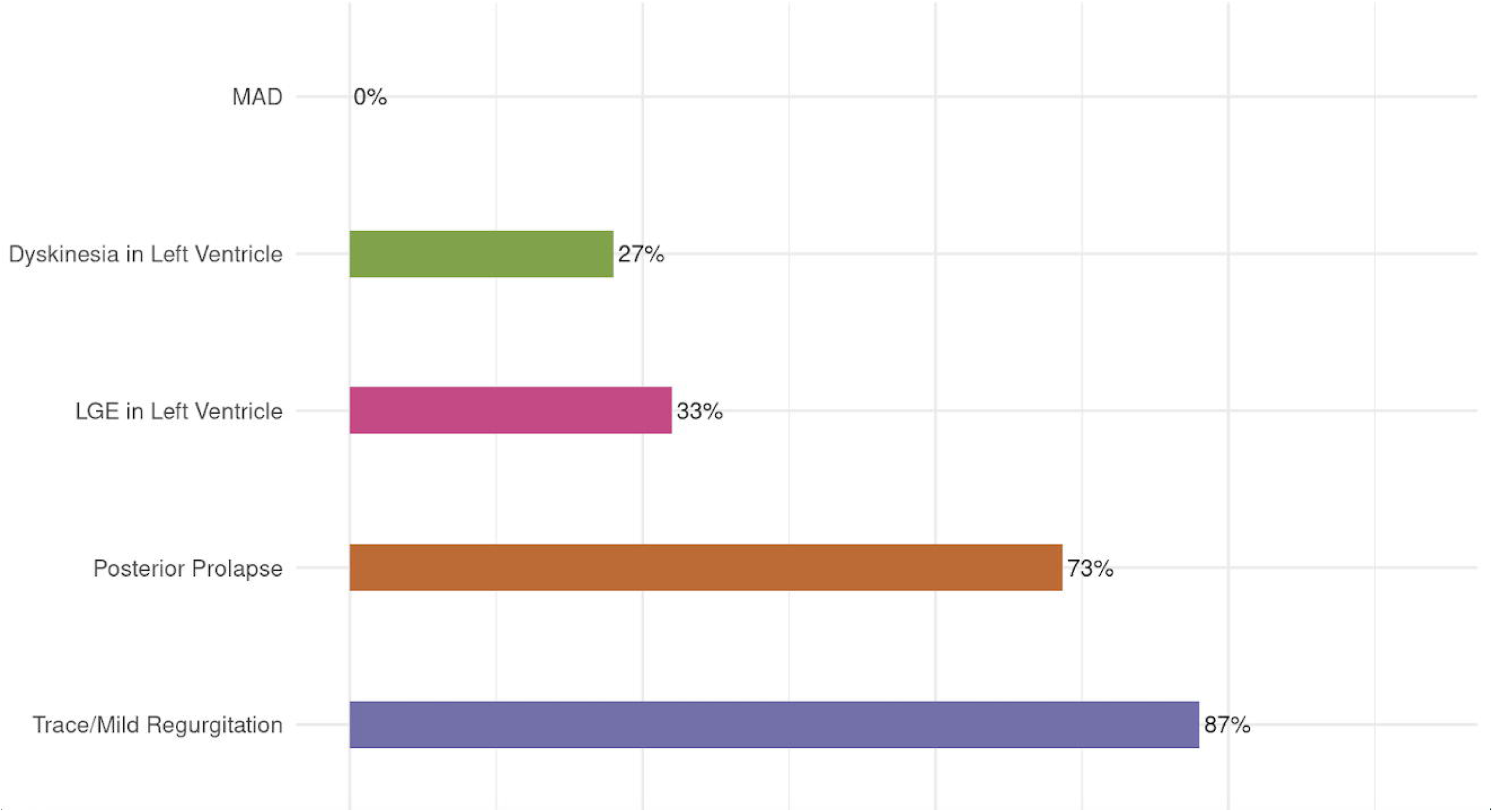

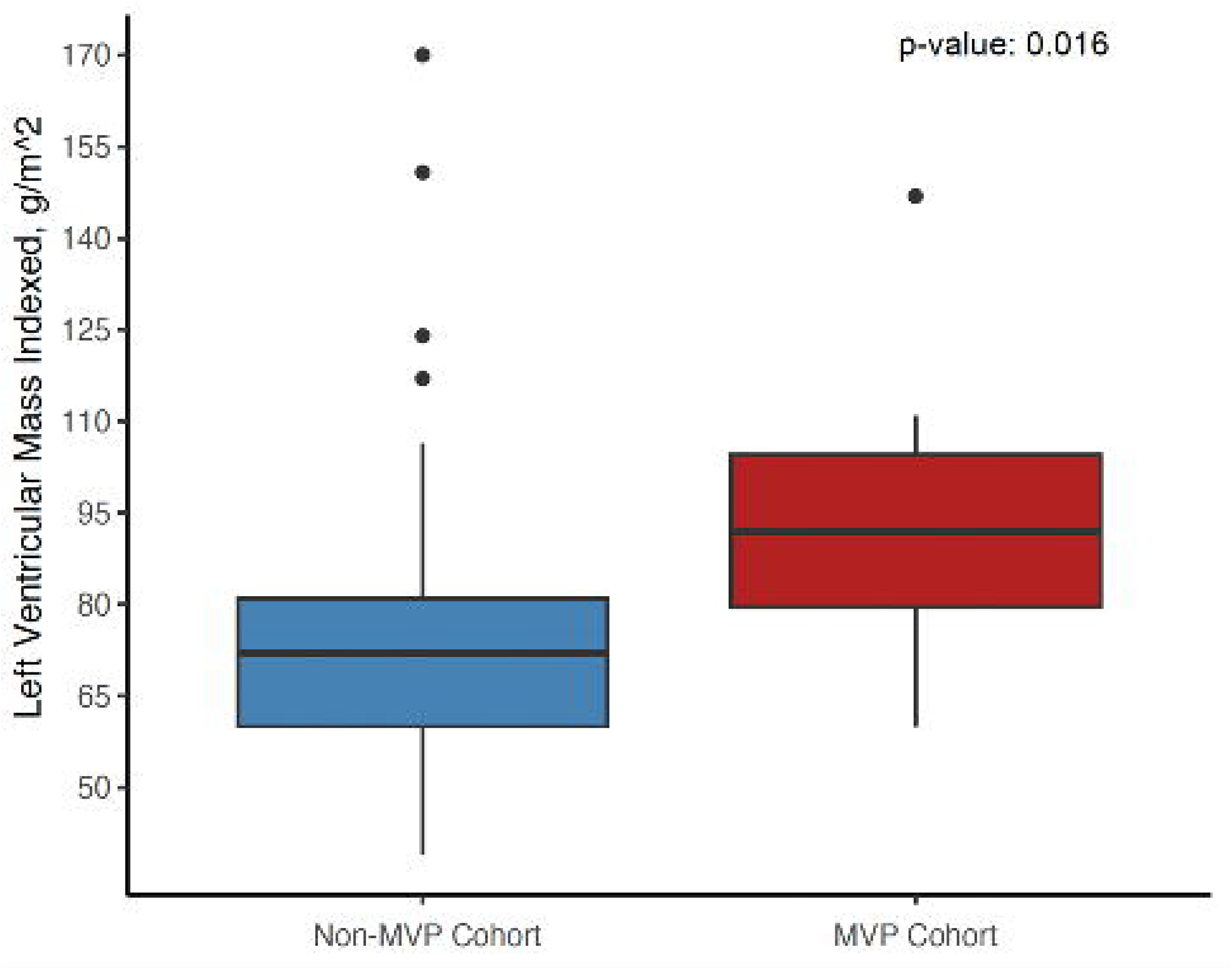

