## Supplemental Material for "Prevalence, Genetics, and Imaging Characteristics of Patients with Mitral Valve Prolapse and Arrhythmogenic Right Ventricular Cardiomyopathy"

**Table S1:** 2010 Task Force Criteria for definite ARVC diagnosis

| **Category** | **Major Criteria** | **Minor Criteria** |
| --- | --- | --- |
| **I. Global or regional dysfunction and structural alterations** | **By 2D echocardiography:** Regional RV akinesia, dyskinesia, or aneurysm and one of the following (end diastole): PLAX RVOT ≥32 mm (≥19 mm/m²), PSAX RVOT ≥36 mm (≥21 mm/m²), or fractional area change ≤33%.  **By CMR:** Regional RV akinesia, dyskinesia, or dyssynchronous RV contraction **and** RV end-diastolic volume/BSA ≥110 mL/m² (male) or ≥100 mL/m² (female), or RV ejection fraction ≤40%. | **By 2D echocardiography:** Regional RV akinesia or dyskinesia and one of the following (end diastole): PLAX RVOT 29–<32 mm (16–<19 mm/m²), PSAX RVOT 32–<36 mm (18–<21 mm/m²), or fractional area change >33% to ≤40%.  **By CMR:** Regional RV akinesia, dyskinesia, or dyssynchronous RV contraction **and** RV end-diastolic volume/BSA 100–<110 mL/m² (male) or 90–<100 mL/m² (female), or RV ejection fraction >40% to ≤45%. |
| **II. Tissue characterization of wall** | Residual myocytes <60% by morphometric analysis (or <50% if estimated), with fibrous replacement of the RV free wall myocardium in ≥1 sample, with or without fatty replacement of tissue on endomyocardial biopsy. | Residual myocytes 60–75% by morphometric analysis (or 50–65% if estimated), with fibrous replacement of the RV free wall myocardium in ≥1 sample, with or without fatty replacement of tissue on endomyocardial biopsy. |
| **III. Repolarization abnormalities** | Inverted T waves in right precordial leads (V1–V3) or beyond in individuals >14 years of age, in the absence of complete right bundle-branch block (QRS ≥120 ms). | Inverted T waves in leads V1 and V2 in individuals >14 years of age (in the absence of complete right bundle-branch block), or inverted T waves in leads V4, V5, or V6. |
| **IV. Depolarization/conduction abnormalities** | Epsilon wave (reproducible low-amplitude signals between the end of the QRS complex and the onset of the T wave) in right precordial leads (V1–V3). | Late potentials by signal-averaged ECG present in ≥1 of 3 parameters in the absence of a QRS duration ≥110 ms on standard ECG; terminal activation duration of QRS ≥55 ms measured from the nadir of the S wave to the end of the QRS (including R′) in V1, V2, or V3, in the absence of complete right bundle-branch block. |
| **V. Arrhythmias** | Non-sustained or sustained ventricular tachycardia of left bundle-branch block morphology with superior axis (negative or indeterminate QRS in leads II, III, and aVF and positive in lead aVL). | Non-sustained or sustained ventricular tachycardia of RV outflow tract configuration, left bundle-branch block morphology with inferior axis or of unknown axis; >500 ventricular extrasystoles per 24 hours on Holter monitoring. |
| **VI. Family history** | ARVC/D confirmed in a first-degree relative who meets current Task Force criteria; ARVC/D confirmed pathologically at autopsy or surgery in a first-degree relative; identification of a pathogenic mutation categorized as associated or probably associated with ARVC/D in the patient under evaluation. | History of ARVC/D in a first-degree relative in whom it is not possible or practical to determine whether the family member meets current Task Force criteria; premature sudden death (<35 years of age) due to suspected ARVC/D in a first-degree relative; ARVC/D confirmed pathologically or by current Task Force criteria in a second-degree relative. |

**Table S1:** Definite ARVC Diagnostic terminology for revised 2010 Task Force Criteria: 2 major or 1 major and 2 minor criteria or 4 minor from different categories.^19^

**Table S2:** Identified genetic variants among ARVC patients

| **Gene** | **HGVS** | **ACMG** | **Cohort** | **Frequency** |
| --- | --- | --- | --- | --- |
| ABCC9 | c.1455T>A  p.(=) | VUS |  | 1 |
| AGL | c.325G>T  p.Val109Leu | VUS |  | 1 |
| AGL | c.1078C>T  p.His360Tyr | VUS(LP) |  | 1 |
| AKAP9 | c.6330+3A>G  p.? | VUS |  | 1 |
| ALMS1 | c.4207A>G  p.Thr1403Ala | VUS |  | 1 |
| ANK2 | c.8892C>G  p.Ile2964Met | VUS |  | 1 |
| BAG3 | c.943T>A  p.Ser315Thr | VUS |  | 1 |
| CALR3 | c.31A>G  p.Ile11Val | VUS |  | 1 |
| DES | c.391C>A  p.Gln131Lys | VUS |  | **2** |
| DES | c.736-8C>A  p.? | VUS |  | 1 |
| DMD | c.7501G>T  p.Gly2501Cys | VUS |  | 1 |
| DMD | c.3986C>A  p.Thr1329Asn | VUS |  | 1 |
| DMD | c.1988C>T  p.Ala663Val | VUS |  | 1 |
| DSC2 | 2326 A>G  p.? | LB |  | 1 |
| DSC2 | c.729del  p.Phe243Leufs*38 | P |  | 1 |
| DSG2 | c.3509_3062delAGAG  p.Glu1020AlafsX18 | P |  | 1 |
| DSG2 | c.1003A>G  p.Thr335Ala | VUS |  | 1 |
| DSG2 | c.2358delA  p.Asp787fs | VUS(LP) |  | 1 |
| DSP | c.4372 C>G  p.Arg1458Gly | VUS(LB) |  | 1 |
| DSP | c.8467C>G  p.Pro2823Ala | VUS |  | 1 |
| DSP | c.2644G>T  p.Glu882* | P |  | 1 |
| DSP | c.5513G>A  p.Arg1838His | VUS |  | 1 |
| DSP | c.273+1G>A  p.? | P | MVP | 1 |
| DSP | c.2683T>C  p.Tyr895His | VUS |  | 1 |
| DSP | c.478C>T  p.Arg160* | P |  | 1 |
| DSP | c.3805C>T  p.Arg1269* | P |  | 1 |
| DSP | c.6516delT  p.F2172Lfs*15 | P |  | 1 |
| DSP | c.2422C>T  p.Arg808Cys | P |  | 1 |
| DSP | c.943C>T  p.Arg315Cys | VUS |  | 1 |
| DSP | c.521G>T  p.Cys174Phe | VUS |  | 1 |
| DSP | c.1556C>T  p.Ala519Val | VUS |  | 1 |
| DSP | c.5213 G>A  p.? | LB |  | 1 |
| FLNC | c.731A>G  p.Asn244Ser | VUS |  | 1 |
| GAA | c.1958C>A  p.Thr653Asn | P |  | 1 |
| GATA5 | c.?  p.? | VUS |  | 1 |
| GATAD1 | c.170G>C  p.Gly57Ala | VUS |  | 1 |
| KCNQ1 | c.239G>C  p.Gly80Ala | VUS | MVP | 1 |
| KCNQ1 | c.1189C>T  p.Arg397Trp | VUS |  | 1 |
| KCNQ1 | c.584G>A  p.Arg195Gln | VUS |  | 1 |
| LAMA4 | c.5185+1G>A  p.? | VUS |  | 1 |
| LAMA4 | c.2594C>T  p.Pro865Leu | VUS |  | 1 |
| LMNA | c.1634G>A  p.Arg545His | VUS |  | 1 |
| MYBPC3 | c.598A>G  p.Ser200Gly | VUS |  | 1 |
| MYBPC3 | c.2504_2505delinsTT  p.Arg835Leu | VUS |  | 1 |
| MYH6 | c.2383C>T  p.Arg795Trp | VUS |  | 1 |
| MYH7 | c.3094G>A  p.Asp1032Asn | VUS(LP) |  | 1 |
| MYPN | c.625T>G  p.Ser209Ala | VUS |  | 1 |
| NEBL | c.903+1G>A  p.? | VUS |  | 1 |
| NEXN | c.856C>T  p.Arg286Trp | VUS(LB) |  | 1 |
| PKP2 | c.235C>T  p.Arg79* | P |  | 1 |
| PKP2 | c.2117T>C  p.Leu706Pro | VUS |  | 1 |
| PKP2 | c.213+3C>G  p.? | VUS |  | 1 |
| PKP2 | c.1613G>A  p.Trp538* | P |  | 1 |
| PKP2 | c.235C>T  p.Arg79* | P | MVP | 1 |
| PKP2 | c.2146-1G>C  p.? | P |  | **8** |
| PKP2 | c.922+1G>A  p.? | P |  | 1 |
| PKP2 | c.2312_2313delTC  p.Leu771ProfsX2 | P |  | 1 |
| PKP2 | c.270_273delinsTGGTTGTAGATGATT p.H91fsX94 | P |  | 1 |
| PKP2 | c.413_586dup  p.? | P |  | 1 |
| PKP2 | c.2197_2202delinsG  p.His733fs*8 | P | MVP | 1 |
| PKP2 | c.1_171del  p.? | P | MVP | 1 |
| PKP2 | c.1034+4del  p.? | VUS |  | 1 |
| PKP2 | c.623del  p.Thr208Lysfs*55 | P |  | 1 |
| PKP2 | c.1803del  p.Asp601Glufs*55 | P |  | 1 |
| PKP2 | c.658C>T  p.Gln220* | P |  | 1 |
| PKP2 | c.517C>T  p.Gln173* | P | MVP | 1 |
| PKP2 | c.808C>T  p.Gln270* | P |  | 1 |
| PKP2 | c.1237C>T  p.Arg413* | P |  | 1 |
| PKP2 | c.1688+1G>A  p.? | LP |  | 1 |
| PKP2 | c.1613G>A  p.Trp538* | P |  | 1 |
| PKP2 | c.968_971delinsGCT  p.Gln323Argfs*29 | P |  | 1 |
| PKP2 | c.2197_2202delinsG  p.His733Profs*8 | P |  | 1 |
| PKP2 | c.2146-1G>C  p.? | P |  | 1 |
| PKP2 | c.1125_1132del  p.Phe376Alafs*8 | P |  | 1 |
| PKP2 | c.466+1A>G  p.? | P |  | 1 |
| PKP2 | c.?  p.? | P |  | 1 |
| PKP2 | c.224-3 C>G  p.? | LP |  | 1 |
| PKP2 | c.235 C>T  p.? | P |  | 1 |
| PKP2 | c.1613G>A  p.W538* | P |  | 1 |
| PKP2 | c.200-4G>C  p.? | VUS | MVP | 1 |
| PKP2 | c.586C>T  p.Arg196Cys | VUS | MVP | 1 |
| PKP2 | c.922+1G>A  p.Asp26Asn | LP |  | 1 |
| PKP2 | c.1097T>V  p.? | LB |  | 1 |
| RYR2 | c.3320C>T  p.Thr1107Met | VUS |  | 1 |
| RYR2 | c.7511C>T  p.Thr2504Met | VUS | MVP | 1 |
| RYR2 | c.11120G>A  p.Gly3707Asp | VUS |  | 1 |
| RYR2 | c.1822C>T  p.His608Tyr | VUS |  | 1 |
| RYR2 | c.1135G>A  p.Val379Met | VUS |  | 1 |
| SAMD9L | c.?  p.? | VUS |  |  |
| SCN10A | c.2130T>A  p.(=) | VUS |  | 1 |
| SDHA | c.?  p.? | VUS |  |  |
| SLC22A5 | c.1043T>C  p.Ile348Thr | VUS |  | 1 |
| SOS2 | c.932A>C  p.Lys311Thr | VUS |  | 1 |
| SPRED1 | c.1320_1321del  p.His440Glnfs*23 | VUS |  | 1 |
| TMEM43 | c. 1073 C>T  p.Ser358Leu | P |  | 1 |
| TRPM4 | c.2479G>A  p.Glu827Lys | VUS |  | 1 |

**Table S2:** ACMG = American College of Medical Genetics and Genomics (classification standard); HGVS = Human Genome Variation Society (nomenclature standard); LB = likely benign; LP = likely pathogenic; MVP = mitral valve prolapse; P = pathogenic; VUS = variant of unknown significance

**CMR Acquisition and Image Analysis**

CMR Acquisition

Acquisition Imaging was performed using the UCSF Core using 3.0T scanners Discovery MR750w or SIGNA Premier (GE Healthcare, Milwaukee, WI). Briefly, standard long- and short-axis cine images were obtained using a balanced steady-state free precession sequence. Typical parameters were TR/TE 2.8/1.3ms, flip angle 45º. Late gadolinium enhancement was performed 5-15 min following injection of 0.1 mmol/kg gadobutrol, and images were obtained using a high-resolution breath-hold two-dimensional sequence at three separate levels in the short-axis plane (basal, mid, and apical). An inversion-recovery fast gradient-echo sequence was performed in two phase-encoding directions to differentiate true late enhancement from artifact (44,45). The inversion time was optimized to achieve satisfactory nulling of the myocardium. T1-weighted imaging with and without fat suppression was utilized to detect fatty infiltration.

Image Analysis

Image analysis was performed offline using a standardized approach (cvi42 version 5.14.0, Circle Cardiovascular Imaging Inc., Calgary, Canada). RV and LV volume and mass were indexed to body surface area. Papillary muscles were included when measuring RV and LV mass and excluded when measuring volumes (45). Areas of inversion artefact or signal contamination by epicardial fat or blood pool were manually excluded. Presence and localization of late gadolinium enhancement were assessed qualitatively. Contrast CMR was performed in selected individuals based on clinical indications using a 3-T magnetic resonance imaging scanner (Discovery MR750w, General Electric Healthcare, Milwaukee, WI, USA). At 10 minutes after injection of 0.1 mmol/kg gadobutrol, late gadolinium enhancement images were obtained with a high-resolution breath-hold 2-dimensional sequence at three separate levels in the short axis plane.

**Genetics and Imaging Characteristics of concomitant MVP and ARVC, excluding borderline MVP cases**

There were 7 diagnostic MVP cases in our ARVC cohort, 6 with available genetic testing and one (17%) with a pathogenic PKP2 variant. Most diagnostic MVPs had monoleaflet involvement (3 posterior, 3 anterior, 1 bileaflet) and trace/mild MR (71%). By CMR, 2 (29%) had LV dyskinesis and 3 (43%) had LV LGE. Comparative analysis of LV mass indexed to body surface area between ARVC patients with (n= 7) versus without diagnostic MVP (n = 104) did not reveal significant differences between the 2 groups (110 g/m^2^ vs. 75 g/m^2^, p = 0.07 by Welch’s t-test). There was a higher prevalence of LV wall motion abnormalities among ARVC patients with diagnostic MVP (43% vs. 6%, p = 0.01 by Fisher’s Exact test.
